# Profile of patients treated with intravitreal anti-vascular endothelial growth factor injections in Bhutan: a 3-year national survey

**DOI:** 10.1101/2022.12.01.22283009

**Authors:** Bhim B. Rai, Deepa Rai, Ted Maddess

## Abstract

**Purpose:** Ocular vascular diseases are common causes of visual impairment and blindness, for which intravitreal injection of anti-vascular endothelial growth factor (anti-VEGF) is the first-line therapy. Current study describes the profile of patients receiving intravitreal anti-VEGF injections in Bhutan. This is the first study of its kind to inform the national health policy.

**Methods:** For this retrospective study, we reviewed the surgical registers of the vitreoretinal unit across Bhutan over three years. Patient demography, clinical findings, diagnostic tests performed, and diagnoses or indications for intravitreal anti-VEGF injections were logged. A descriptive analysis was performed.

**Results:** A total of 381 patients received intravitreal anti-VEGF injections in the operating theatres as mandated by the national guidelines. The majority of patients were males (230, 60.4%). The mean age was 65.2 ± 13.5 years, ranging from 13 to 90 years, and a median of 69 years. Majority of the treated eyes (117, 30.7%) had BCVA <3/60 to light perception (PL), and another 51 eyes (13.4%) had < 6/60 to 3/60. The most common indication for anti-VEGF injection was neovascular AMD (168 cases, 42.2%), followed by retinal vein occlusion (132 cases, 34.6%), diabetic macular oedema and retinopathy (50 cases, 13.1%), and myopic choroidal neovascular membrane (11 cases, 0.03%).

**Conclusions:** Bhutan faces both economic and geographic challenges, on top of limited human resources for managing vitreoretinal diseases. With an ever-increasing load of vitreoretinal diseases, and systemic diseases like diabetes and hypertension, there is a need to improve vitreoretinal services. Regular vitreoretinal services are provided only at the national referral hospital located in the north-west. For successful management, an effective community screening program, right referrals, and proper transport facilities must go hand-in-hand, and or extend regular vitreoretinal services to regional referral hospitals.

## INTRODUCTION

Vascular diseases such as neovascular age-related macular degeneration (nAMD), diabetic macular oedema (DMO) and diabetic retinopathy (DR), and retinal vein occlusion (RVO) are the major causes of visual impairment and loss of central vision throughout the world (1). Previously refractory cases were treated with intravitreal injection of corticosteroid triamcinolone acetonide and dexamethasone (2). However, these agents were associated with multiple ocular side effects such as cataract and intraocular pressure spikes (2). The pathogeneses of many such diseases are multifactorial. Both inflammatory mediators and vasogenic mediators, such as vascular endothelial growth factors (VEGFs), are activated and create disruption of the retina (3, 4). As the inner blood-retinal barrier breaks down, fluids and proteins leak into the retina. This leakage creates oedema of the macula and thereby decreases the retinal transparency (2-5). To counter this effect, anti-VEGFs came into clinical use with much promise. It also earned the name of a *one day wonder drug* for the fast resolution of DMO following intravitreal injection of ranibizumab (6). Among anti-VEGFs, bevacizumab (AVASTIN, Genentech, Inc.,) was the first agent to be used off-label in 2005. Subsequently, multiple anti-VEGF agents such as pegaptanib sodium (Eyestech/OSI Pharmaceuticals, New York, NY, USA), ranibizumab (Lucentis™, Genentech, Inc., South San Francisco, CA, USA and Novartis Pharma AG, Basel, Switzerland), aflibercept (VEGF-trap eye, Eylea™, Regeneron Pharmaceuticals, Inc., and Bayer Pharma AG, Berlin, Germany), brolucizumab (Beovu™, Novartis Pharma AG, Basel, Switzerland), and faricimab have been developed for the management of various ocular vascular diseases (7-11). Although all these agents are used, only pegabtanib, ranibizumab, brolucizumab, aflibercept, and faricimab are approved for use in ophthalmological disorders (11-13). Ranibizumab, bevacizumab and brolucizumab specifically target VEGF-A activity, while aflibercept has a broader spectrum of action – VEGF-A, as well as other VEGF family ligands such as VEGF-B, placental growth factor-1 (PlGF-1) and placental growth factor-2 (PlGF-2) (14, 15). Faricimab is a new and the first bispecific monoclonal antibody for intravitreal use neutralising both VEGF and angiopoietin-2/tie-2/ pathway(16). Due to the prolonged activity, faricimab allows extended intervals between injections of up to three or four months in nAMD and DMO patients, which can be a significant benefit for patients and are an alternative to implanted drug delivery systems (17, 18). The aim of the current study was to report on the profile of patients treated with intravitreal anti-VEGFs injections for ophthalmological diseases in Bhutan. Since this is the first study of this kind in the country and the results have impacts for policy making in Bhutan and other developing countries in the region.

## METHODS

### Study design and Ethics

This was a retrospective, case series study approved by the Research Ethics Board of Health (REBH) (REBH/Approval/2016/083), Ministry of Health, Royal Government of Bhutan, Thimphu, Bhutan. This study adhered to the principles of the Declaration of Helsinki. Informed consent was waived by REBH because this retrospective study collected only de-identified data.

### Setting

In Bhutan the Jigme Dorji Wangchuck (JDW) national referral hospital is the only apex national referral and teaching hospital with subspecialty eye-care services. Regular vitreoretinal surgical and interventional services in the country are provided only at the JDW national referral hospital. All such vitreoretinal patients across the country are referred there for management. Occasionally the vitreoretinal surgeon visits Eastern Regional Referral Hospital (ERRH) in Mongar and Central Regional Referral Hospital (CRRH) in Gelephu when the treatment is provided there. The sites for the current study included JDW national referral hospital (in the main), and ERRH and CRRH.

### Participants

All the patients who received intravitreal anti-VEGF injections in the operation theatres at the JDW national referral hospital, ERRH and CRRH over three years (01 May 2013 until 30 April 2016) were included. The national guidelines mandate that intravitreal injections should be administered only in operating theatres with proper aseptic measures. The demographic characteristics of patients receiving the anti-VEGF injection and the vitreoretinal disease pattern represent the entire citizenry.

### Clinical Examination and data collection

The clinical workup in the out-patient department (OPD) included taking detailed medical histories of presenting symptoms and diseases, associated systemic diseases, and prior surgical or laser interventions done. Best corrected visual acuity (BCVA) at presentation was measured using a Snellen chart, and Tumbling E for illiterate patients. Intraocular pressure was measured by Goldmann applanation tonometry or an iCare tonometer, and the anterior and posterior segments were examined under slit-lamp biomicroscopy (BM 900, Haag-Streit, Switzerland) and 90D bio-microscopy. The funduscopic findings were confirmed by binocular indirect ophthalmoscope (Model 125, Welch Allyn, USA). Macular and retinal nerve fibre (RNFL) scans were measured using a Spectral Domain Optical Coherence Tomography (OCT) (Cirrus-HD 4000, Carl Zeiss Germany). Fundus photographs were taken by a VISUCAM-524 (Carl Zeiss, Germany). Fundus fluorescein angiograms (FFA) were performed when needed to confirm occult diseases. A B-ultra sonogram scan was done when necessary (Sonomed Escalon, Model: VUPAD A/B, USA).

For the current study, the patient register maintained in the vitreoretinal operational theatre was reviewed for the data collection. The data collected included demographic information such as age and gender, residential settings, BCVA, diagnostic tests performed, and diagnosis or indications for anti-VEGF injections.

### Diagnostic tests

OCT was performed universally on all cases who have received intravitreal anti-VEGF injections. This was an appropriate diagnostic tool to diagnose nAMD, RVO, DMO, myopic choroidal neovascular membrane (mCNVM), cystoid macular oedema, central serous chorioretinopathy and vitreomacular tractions. Polypoidal choroidal vasculopathy (PCV) is best diagnosed via its characteristic nodular dilatations arising from neovascular networks that ramify mainly in the subretinal pigment epithelial space on indocyanine green angiography.(19, 20) However, in the current study the PCV was mainly diagnosed based on the OCT findings such as sharp-peaked retinal pigment epithelial detachment (RPED), subretinal pigment detachment ring-like lesion, multilobular RPED, double-layer sign or shallow, irregular <u>retinal pigment</u> epithelium elevation, thick choroid with dilated Haller’s layer vessels, and predominance of subretinal fluid. Other suggestive findings with the coloured fundus photographs were extensive subretinal haemorrhage, and orange nodules.(21) The FFA was performed only in difficult or occult cases.

### Anti-VEGF injections

Most of the time, a 4 ml vial containing 100 mg bevacizumab was procured. The patients needing intravitreal anti-VEGF injections were pooled for a scheduled injection day, when a vial was opened and dosing formulation done at 1.25 mg per 0.05 ml, or 2.5 mg per 0.1 ml in tuberculin syringes with 30G needles. So a single vial would benefit 20 to 25 pooled patients. This approach was practised for economic reasons. The anti-VEGF dosing formulations and intravitreal injections were performed under aseptic measures in the operating theatres. Injections into the bilateral eyes in the same siting was not performed to avoid complications, notably endophthalmitis.

## RESULTS

### Demographic characteristics

A total of 381 patients received intravitreal anti-VEGF injections over the study period. The majority of patients (230, 60.4%) were males, and others females (151, 39.6%). The mean age at presentation was 65.2 ± 13.5 years (ranging from 13 to 90 years), modal year of 70 years, and median 69 years.

### Visual acuity

BCVA at presentation was recorded in the vitreoretinal unit OPD register and the patient personal files. For the current study we referred to the surgical register maintained in the operating theatre, where BCVA for all cases were not recorded. As a result, the data on BCVA was not available (NA) in 85 cases (Table 1). Among those with BCVA data available, majority of the eyes treated with anti-VEGF injections had BCVA of less than 3/60 to only perception of light (PL), labelled as blindness according to World Health Organisation (WHO) classification of visual impairment, in the treated eyes (117 cases, 30.7) (22). We could not ascertain if they were legally blind with this because we did not have the data on BCVA of their fellow eyes. The details of the BCVA are shown in **Table 1**.

**Table 1.** Best corrected visual acuity at presentation

| *Visual acuity based on WHO*<br>*classification of blindness* | *No. of*<br>*eyes* | *Percentage* |
| --- | --- | --- |
| $\geq 6/18$ | 21 | 5.5 |
| $< 6/18 - 6/60$ | 107 | 28.1 |
| $< 6/60 - 3/60$ | 51 | 13.4 |
| $< 3/60 - PL^*$ | 117 | 30.7 |
| NA<sup>†</sup> | 85 | 22.3 |
| NPL<sup>^</sup> | 0 | 0 |
| Total | 381 | 100 |

NA†: Data not available; NPL^: No perception of light; PL\*: Perception of light.

### Intravitreal anti-VEGF injections

nAMD was the most common indication for the intravitreal anti-VEGF injection, accounted for 168 cases (44.2%). This was followed by RVO in 132 cases (34.6%). Within the RVO subgroup, branch RVO (BRVO) was the most common subgroup with 70 cases (53.0%), followed by central RVO (CRVO) in 55 cases (41.7%), and hemi-retinal BRVO (HRBRVO) in 7 cases (5.3%). Only 50 cases (13.1%) receiving anti-VEGF injections had DR or DMO, 11 cases (0.03%) had mCNVM, and 6 cases (0.02%) had PCV. Other indications for anti-VEGF injections are shown in **Table 2**.

**Table 2:** Indications of anti-VEGF Injection.

| *Indications* | *Number of cases* | *Percentage* |
| --- | --- | --- |
| *Neovascular AMD* | 168 | 44.2 |
| *Total Retinal Vein Occlusion* | 132 | 34.6 |
| *Branch Retinal Vein Occlusion (BRVO)* | 70 | 53.0 |
| *Central Retinal Vein Occlusion (CRVO)* | 55 | 41.7 |
| *Hemi-retinal BRVO (HRBRVO)* | 7 | 5.3 |
| *Diabetic Macular Oedema* | 50 | 13.1 |
| *Myopic CNVM (mCNVM)* | 11 | 0.03 |
| *Polypoidal Choroidal Vasculopathy (PCV)* | 6 | 0.02 |
| *Neovascular Glaucoma* | 4 | 0.01 |
| *Others\** | 10 | 0.03 |
| *Total* | **381** | **100.0** |

2 cases of injection were excluded as the indications were not recorded.
\*Others include Cystoid macular oedema, Central serous chorioretinopathy and Vitreomacular traction

## DISCUSSION

To summarise the vital results, 30.7% of the study eyes were legally blind, 13.4% had severe visual impairment (VI) and 28.1% has moderate VI as per the WHO classification of VI (22). nAMD was the most common indication for intravitreal anti-VEGF injections, followed by RVO, DMO/DR, mCNVM, PCV, and neovascular glaucoma.

A study on the pattern and presentation of vitreoretinal diseases in Bhutan in the same period reported that majority of patients were males, 1544/2913 (23). That is somewhat lower than the rate seen here (p=0.048, χ^2^=3.90). A meta-analysis of eye surgical services in developing countries has shown that females receive fewer surgical or interventional services than the males (24), as was seen with receiving anti-VEGF injections in the current study. The national vitreoretinal unit was established in Bhutan only in early 2012 (25), Prior to that, vitreoretinal cases were referred to India or Nepal for treatment. Most patients were reluctant to go out of the country for surgery, however, because of the communication gap in foreign lands. This partly encouraged some patients to seek to traditional or indigenous methods of healing. Some even sought to local healers or witchcraft practice with false hope, resulting in irreversible loss of vision and even loss of eyes, while most cases did not receive any treatment and accepted it as their destiny.

AMD was reported as the fourth most common vitreoretinal disease in Bhutan, next to hypertensive retinopathy, refractive errors referred for retinal evaluation, and DR or clinically significant macular oedema (CSMO) (23). So it is apparent that intravitreal anti-VEGF injection was most commonly administered to nAMD cases. Another study on the profile of AMD in Bhutan reported that half of AMD cases were in advance stages needing treatment, and one-third of those with advanced diseases already had disciform scar at their first presentation (26). Similarly, the drop-out rates for follow-up are high. The reason for such late presentation and poor follow-up or medical adherence are many. First, lack of knowledge of the general public on diseases and treatment options available. Secondly, lack of accessibility to services – the regular vitreoretinal service is provided only at the JDW national referral hospital located in the north-west part of the country and the patients from the central and eastern regions have to travel all the way against difficult terrain, poor road conditions and transport limitations to avail the treatment. Other reasons include poor referral system, communication gap between the peripheral and referral hospitals, and negative influence of the local healers.

Co-existent hypertension and diabetes mellitus, the complications of which result to many ocular vascular diseases, is common in Bhutan (27). WHO has reported that 28% of total mortality in Bhutan was due to cardiovascular disease, and another 17% due to other non-communicable diseases (28). For these morbidities, the risk factors include harmful use of alcohol, high salt intake, unhealthy diet, physical inactivity, and tobacco use (29). Considering hypertensive retinopathy and diabetes mellitus as risk factors for RVO (30), and both being common among the Bhutanese population, RVO cases received the second highest number of anti-VEGF injections. Similarly, RVO cases also was the second most common indication to receive retinal laser therapy in the country, second only to DR/DMO (31). Only 50 cases (13.1% of all anti-VEGF injections) received anti-VEGF injection in the study population although DR and DMO was the third most common vitreoretinal disease in Bhutan, and second most common in a similar study conducted in Nepal (32). This is because only the central DMO, and DMO nasal to the fovea, were treated with anti-VEGF injection. Very severe DR not responding to panretinal photocoagulation (PRP) were also first treated with the anti-VEGF injections to reduce retinal oedema after which PRP was performed successfully. Commonly, the DR cases were managed with PRP to such a rate that PRP was the most common type of retinal laser therapy performed in Bhutan (31). Off-centre DMO (except DMO nasal to the fovea) was managed with focal, grid or modified grid laser therapies. An unpublished study (under review with the Japanese J of Ophthalmology) has reported that 25.6% of diabetes patients presented 5-10 years after diagnosis, 19.9% after 10-20 years and 2.3% presented only after 20 years for the first time for their eye/retinal check-up. The study also reported that 12.5% of DR cases had proliferative DR at their first presentation for retinal evaluation.

Although the moderate to high myopic cases referred for retinal evaluation was the second most common reason for attending vitreoretinal clinics in Bhutan, next to hypertensive retinopathy (23), only a small percentage of anti-VEGF injections was given to cases of myopic CNVM (mCNVM). This is because the prevalence of mCNVM among the study group of myopia was only 0.4% (33). Other studies have reported that mCNVM occurs in approximately 5-10% of highly myopic patients, and 0.04-0.05% of the general population (34, 35). PCV is common chorioretinal disorder among the Asian populations.(36) It has been estimated that up to 50% of nAMD cases among the Asians are of the PCV subtype, whereas this proportion among the white populations is estimated to range between 10% and 20% (37, 38). In Bhutan, 8.1% of advanced AMD were comprised of PCV, and among them 73.7% of them (14/19 cases) were younger than 50 years (26). This is highly concerning for a country like Bhutan with a small reserve of working age population and limited human resources. Yet only six of nineteen cases of PCV received anti-VEGF treatment, that is, 68.4% of them did not receive treatment. There is fear that they may turn up later with worsened disease not being amenable to the treatment. With these concerns, it was also deliberated that community screening of AMD along with PCV cases might be helpful in Bhutan (39). However, until today, it seems there are no large-scale screening programs for AMD like those that exist for DR and retinopathy of prematurity.

It is common that younger patients have poor medical adherence (40). So the treating physicians and the healthcare professionals have to give extra effort to teach them about the disease and ensure the importance of timely medications. Additionally, the general population in Bhutan are illiterate and have poor knowledge of diseases and treatment options. Although health education is provided at all levels of healthcare centres in Bhutan there is need to improve the program. It may be effective to disseminate health advocacy information in the local languages or dialects so the common people are able to understand the message. Involving local leaders and local government bodies may prove more effective. Broadcasting the materials through the national television channels, or even targeting the younger generations through the mobile phone applications they use, may be helpful. For regionally balanced delivery of the services, Bhutan needs to extend its regular vitreoretinal services to the ERRH and CRRH, or at least improve community screening and effective referral system. New approaches to screening and management of the AMD cases may allow for expansion of high-quality convenient care and increasing patient coverage (41). The public transport system in Bhutan is challenged by difficult terrain, poor quality of roads and infrequent services. Additionally, it is interrupted during monsoon season by landslides, falling boulders and swollen rivers and streams, and by snowfall during winters. Therefore, the proper arrangement of transport facilities for the patients is a must for successful screening and referral program. For referrals too, a concept of *right referral* must be practised. Only the patients needing to be seen by the retinal specialist and interventions at the national referral hospital should be referred. If patients are referred only to be told that no further action required after taking a difficult journey, then such improper referral will certainly have negative impact on the program. Tactfully, Bhutan also needs to lower the influence of local healers and encourage people to visit hospitals while having health issues. Although Bhutan still struggles to provide basic medical provision and low-cost treatment, it may be worth to identify patients who are not compliant to follow up or adherent to medication and provide longer acting anti-VEGF treatment such as faricimab, which can extend the intervals between successive drug administration by 4 to 6 months (17, 18).

This study is limited by being a retrospective study and a small number of study subjects. We did not have visual acuity at presentation for 85 cases (22.3%). Being a cross-sectional study, we also did not follow up the patients to understand the outcome of the anti-VEGF treatment. We have a plan to conduct a prospective study overcoming all the limitations of the current study.

## CONCLUSIONS

There is need to improve the vitreoretinal services in Bhutan. Currently regular vitreoretinal service is provided only at the JDW national referral hospital, so there is high risk that the patients from the eastern and central regions miss out on the treatment and regular follow up. So regular services need to be expanded to the eastern and central regional referral hospitals. Secondly, vitreoretinal diseases secondary to systemic diseases such as hypertension and diabetes are common. There is an urgent need for strategic control and management of non-communicable diseases and prevent blindness and visual impairment from their complications.

## Data Availability

The data is available with the corresponding author upon request.

## DECLARATIONS

### Conflict of interest

Author BBR declares that he has no conflict of interest. Author DR declares that she has no conflict of interest. Author TM declares that he has no conflict of interest.

### Funding

The funders had no role in study design, data collection and analysis, decision to publish, or preparation of the manuscript.

### Ethical approval, Informed Consent and Consent for Publication

The study was approved by the Research Ethics Board of Health (REBH) (REBH/Approval/2016/083), Ministry of Health, Royal Government of Bhutan, and was conducted as per the REBH guidelines and tenets of the Declaration of Helsinki. The consent was waived by REBH as the retrospective study collected only the de-identified data.

### Availability of data and material

The data and material are available with the corresponding author.

## Notes

### Competing Interest Statement

The authors have declared no competing interest.

